# Validation of the Food Safe Zone Questionnaire for Families of Individuals with Prader-Willi syndrome

**DOI:** 10.1101/2024.01.26.24301844

**Authors:** Elisabeth M Dykens, Elizabeth Roof, Hailee Hunt-Hawkins

**Affiliations:** Vanderbilt University, Department of Psychology and Human Development, Nashville, TN, U.S.A.

**Author notes:** Corresponding Author (EMD).

## Abstract

Prader-Willi syndrome (PWS), a genetic neurodevelopmental disorder, is characterized by hyperphagia and significant behavioral problems. Hyperphagic individuals with PWS are chronically hungry yet rarely feel sated, and often engage in food-seeking behaviors. To avoid obesity in their children, families implement food security and supervision strategies (e.g., locking food sources, alerting others). Yet such accommodations may undermine parental appraisals of hyperphgia (e.g., it’s not a problem as we lock food); a potential confound in clinical trials aimed at attenuating hyperphagia. We developed the Food Safe Zone (FSZ) questionnaire to remind parents of their food safety practices prior to them evaluating their child’s hyperphagia.

Our team developed 20 FSZ items that were revised for clarity and completeness in an iterative feedback process with stakeholders; parents, PWS specialists, and individuals with PWS. The FSZ was pilot tested, descriptive findings were reviewed by additional stakeholders, and then administered to 624 parents in a large-scale study. Based on an open-ended question, “Is there anything else you do to ensure food safety?” two additional items were added and evaluated in a follow-up study.

Principal component analyses revealed that 21 FSZ items loaded onto 5 factors that were readily interpretable, accounting for 67% of test variance: Alerting Others and Food Supervision in the Community; Locking or Restricting Food Sources; Checking for Food; At Home Supervision and Meals; and Avoiding Food Settings. Internal consistency and test-rest reliability were robust. Convergent validity analyses revealed that regardless of age, parents implemented more FSZ strategies in response to the severity of their child’s hyperphagia.

The psychometrically sound FSZ stands to enhance accurate parental assessments of hyperphagia in future studies or clinical trials. Analyses of the open-ended question also shine a light on the extraordinary measures that parents use to ensure the health of their individual with PWS.

## Introduction

Prader-Willi syndrome (PWS) is a neurodevelopmental disorder caused by the lack of paternally imprinted genetic information on chromosome 15q11-q13 [1]. Most cases (~70%) are attributed to paternal deletions in this region that vary in size, or maternal uniparental disomy (mUPD), when the child inherits two copies of the maternal chromosome 15 [2]. Behaviorally, people with PWS typically exhibit salient compulsivity, anxiety, needs for sameness, rigid thinking, hoarding non-food items, temper outbursts, repetitive questioning, and skin-picking [3, 4]. They also have mild to moderate deficits in intellectual and adaptive functioning [5], as well as in social cognition [6], and executive functioning [7, 8].

Hyperphagia, however, is the most striking and distinctive characteristic of PWS. Beginning in early childhood, hyperphagia is attributed to aberrant neural feedback mechanisms involved in appetite regulation and satiety [9]. As a result, hyperphagic individuals with PWS view the world through a lens of hunger; they are constantly hungry, yet rarely feel full or sated [10]. Their unrelenting hunger leads to food seeking behaviors, including sneaking food and manipulating others to obtain food. Hyperphagic severity and behaviors are often assessed with the informant-based Hyperphagia Questionnaire (HQ) [11], and an adapted version of the HQ, the HQ-Clinical Trials (HQ-CT) [12]. The HQ-CT has shown robust responsivity to treatment in previous clinical trials aimed at attenuating hyperphagia and related problems [13, 14,15].

Hyperphagia in PWS is life-threatening as it can lead to medical complications associated with morbid obesity, gastric distention, and necrosis, and choking while binging or sneaking food [16, 17]. Risks of morbid obesity are lessened with growth hormone therapy (GHT), which is now a pediatric standard of care for treating growth hormone deficiencies in PWS. GHT is associated with increased linear height and lean muscle mass, and reduced body fat [18], as well as with advantages in cognition and adaptive behavior [19]. GHT does not, however, effectively treat or lessen the syndrome’s characteristic hyperphagia. To avoid obesity, many families also keep track of the daily caloric content of food consumed by the person with PWS. Doing so is helpful as people with PWS have a lower absolute energy expenditure than others, and thus need fewer calories to lose or maintain a healthy weight [20].

Fortunately, PWS has now garnered the attention of several pharmaceutical companies that are sponsoring clinical trials of novel agents aimed at attenuating hyperphgia and related symptoms in PWS [21]. Although these are promising developments, hyperphagia is currently best managed with environmental food safety strategies [2]. Common food safety strategies include locking food sources (e.g., cabinets, refrigerators, pantries, trash); supervising individuals whenever food is present; coordinating food intake with school, work, or recreational settings; and educating family members, friends, neighbors, teachers, and others about PWS and the need for food restrictions.

Many parents report that over time, their food safety practices become the “new normal” and an integral part of their daily routines. As one parent in our research program observed, “Maintaining food safety has been automatic behavior 24/7 for 30 years.” Automating food safety practices into daily life both facilitates family functioning and helps keep individuals with PWS from becoming dangerously obese.

At the same time, however, successfully implementing food safety practices may lead parents to minimize or underestimate the severity of their child’s hyperphagia, i.e., it’s not a problem because we lock up food, constantly supervise, adhere to a strict meal schedule, etc. Indeed, both the U.S. Food and Drug Administration (FDA) and clinical trial sponsors have raised questions if parental accommodations to their children’s hyperphagic symptoms could undermine their accurate responses on the HQ-CT. Precise measurements of hyperphagia are critical both as an endpoint assessing treatment efficacy in clinical trials and as a criterion for trial eligibility.

In the present study, we developed and validated a novel index of food safety tactics in PWS, the Food Safe Zone (FSZ). The FSZ aims to remind parents of the various strategies they use to manage their child’s hyperphagia prior to them completing the HQ-CT or other measures of hyperphagia. Consistent with the FDA’s guidelines for Observer-Reported and Patient-Reported Outcomes [22], we developed the FSZ by involving multiple stakeholders and adhering to psychometric principles involved in questionnaire development. The study also analyzed the FSZ in relation to demographic variables (e.g., age, gender, PWS genetic subtype, parental socio-economic status). Beyond these analyses, we conducted a content analysis of parental comments to an open-ended question; “What did we miss? Are there other things that you do to ensure food security?” In brief, we anticipate that the FSZ will be an important component of accurately assessing hyperphagia in future research and clinical trials in PWS.

## Methods, Procedures and Participants

### FSZ Item Development and Stakeholder Input

Our research team developed a pool of 20 items that tapped common, readily observable food safety practices. Items were gleaned from interviews, data, and clinical notes from over 325 families enrolled in our previous and current PWS research programs.

Parents of 4 individuals with PWS provided initial feedback on the clarity of items, and the proposed response scale. We also asked them if additional items were needed to fully measure food safety, i.e., construct validity. Parents suggested a rewording of 8 items and nominated 5 additional items on the frequency that individuals with PWS attended events that did or did not involve food, with or without parental supervision.

The revised 25-item FSZ draft was then vetted by 12 parents of individuals with PWS, aged 5 to 33 years, in 4 different, 40-minute focus group sessions held on-line via Zoom. Parents concurred that the wording of the FSZ was clear and did not suggest additional items. Instead, parents specified how they implemented various FSZ strategies. They also mentioned that they are not typically asked about their food safety practices, and that it was helpful to learn how others managed their child’s hyperphagia.

Finally, we solicited input on the FSZ via a Zoom focus group with 5 adults with PWS, aged 20 to 42 years. Group members were forthright about their struggles with hunger and food-seeking. They were asked which FSZ tactics they found helpful, and if we missed any strategies. The group did not suggest new FSZ items, and instead reinforced the importance of securing food, educating others about PWS, and maintaining a predictable meal and snack schedule.

### Pilot Testing and Additional Stakeholder Input

The revised, 25-item FSZ was pilot tested in 133 parents of individuals with PWS aged 5-43 years (see Participants, Table 1). Respondents were asked to complete the questionnaire based on the last month, using the following rating scale: 1 = Never/Rarely; 2 = Some of the time; 3 = Most of the time; 4 = All the time. The FSZ also included an open-ended question, “What did we miss? Are there other things you do to ensure food security?”

**Table 1.** Demographics of participants with PWS and their families for the pilot, large-scale and follow-up studies.

|  | Pilot | Large-Scale | Follow-up |
| --- | --- | --- | --- |
| Demographic Variables | M (SD) or % | M (SD) or % | M (SD) or % |
| N | 133 | 624* | 119 |
| Age, Years | 18.86 (7.89)<br>Range: 5-43 | 18.60 (10.65)<br>Range: 5-59 | 14.25 (7.75)<br>Range: 5-44 |
| Gender | 42.1% Male<br>57.9% Female | 46% Male<br>54% Female | 48% Male<br>52% Female |
| **Genetic Subtypes** |  |  |  |
| Paternal Deletion | 61.1% | 50.7% | 48.8% |
| mUPD | 32.8% | 34.4% | 43.1% |
| Imprinting Deficit | 3.8% | 3.2% | 3.1% |
| Other | 2.3% | 2.3% | 1.3% |
| Unknown |  | 9.4% | 3.8% |
| **Education Maternal/Paternal** |  |  |  |
| High School | 21.4%, 27.0% | 17.2%, 24.4% | 18.8%, 23.3% |
| 2-year college | 16.8%, 10.3% | 29.8%, 30.6% | 15.0%, 11.3% |
| 4-year college | 31.3%, 32.5% | 34.2%, 26.9% | 28.1%, 31.4% |
| Post-graduate | 30.5%, 30.2% | 18.9%, 18.1% | 38.1%, 34.0% |
| **Annual Income** |  |  |  |
| \$10,000 - \$30,000 | 6.1% | 5.8% | 8.8% |
| \$31,000 - \$50,000 | 4.5% | 16.7% | 8.8% |
| \$51,000 - \$70,000 | 13.8% | 16.0% | 13.8% |
| \$71,000 - 100,000 | 20.8% | 17.1% | 16.4% |
| > \$100,000 | 54.6% | 35.3% | 52.2% |
| Unknown | -- | 9.1% | --- |

| **Race, Ethnicity** |  |  |  |
| --- | --- | --- | --- |
| White | 92.3% | 85.2% | 88.7% |
| Hispanic | 3.1% | 7.4% | 5.2% |
| Asian | 2.3% | 2.5% | 3.5% |
| Black | 2.3% | 2.3% | 2.6% |
| Multi-racial |  | 2.6% |  |

Descriptive pilot data (e.g., means, frequencies) were then reviewed by members of FPWR’s Clinical Trials Consortium, specifically 3 parents and 5 PWS professionals. Members concurred that items were clearly worded. They also noted a potential confound with intellectual disability for the parent-nominated questions on letting their individual with PWS attend gatherings with or without food or parental supervision. Nevertheless, the committee recommended retaining these items to determine how they performed in the large-scale study.

### Large-scale Administration

The FSZ was administered to additional parents of individuals with PWS via The Global PWS Patient Registry, a secure, web-based Registry sponsored by the Foundation for Prader-Willi Research (FPWR) and hosted on the National Organization for Rare Disorders “IAMRARE” registry platform [23]. Registrants are asked to complete medical and behavioral questionnaires every six months, a time frame established by FPWR to reduce parental burden of more frequent assessments. The Registry garnered 491 respondents, and 304 (62%) completed the FSZ 6 months later.

### Respondent Feedback and Follow-Up Study

A follow-up study was conducted to assess two additional items gleaned from parental responses to the open-ended question, answered by 78% of respondents. Our team reviewed these responses to determine if they represented novel strategies not included in the FSZ or were instead embellishments or descriptions of how families enacted the tactics already included in the FSZ (see Results). Based on this review, two additional questions were added: “Making a food plan for child prior to attending events, outings, restaurants,” mentioned by 32% of respondents; and “Avoid eating in front of child unless they are also eating,” noted by 37% of respondents. We added these two items to the FSZ and administered the now 27-item FSZ to 119 parents of individuals with PWS in a follow-up study.

### Informed Consent and IRB Approval

Approval for this study was obtained by the Vanderbilt University Institutional Review Board, Integrated Science Committee. Vanderbilt University participants provided written, informed consent using the e-consent function of RedCap, a secure, web-based data collection platform [24]. After consenting, parents were then invited to complete the FSZ, HQ-CT and Demographic questionnaires on RedCap. Additional study approval was obtained for participants recruited from the FPWR Patient Registry. Prior to collecting data from the Registry, the study was reviewed and approved by FPWR’s research committee and internal IRB. All registrants in FPWR’s Patient Registry give approval for their unidentified data to be used for research purposes.

### Participants

Pilot study participants (n=133) were recruited by our team at Vanderbilt University via postings on PWS-related social media platforms, and announcements at regional and national PWS conferences. As previously described, participants for the large-scale study (n=491) were recruited via the FPWR Patient Registry.

Table 1 summarizes demographic variables for participants in the pilot and large-scale studies. No significant differences emerged between these two recruitment sources in participant demographics or FSZ scores. As such, they were combined to form a large group (n=624) to increase power and facilitate statistical analyses.

Participants in the follow-up study (n=119) were recruited from both Vanderbilt University and FPWR (see Table 1). After consenting, they completed the 27-item FSZ, which included the two additional items generated by parent feedback. Approximately 15% of those in the follow-up study had completed the 25-item FSZ one to two years prior either in Vanderbilt’s Pilot study, or FPWR’s Patient Registry.

### Other Measures

#### Demographics

Parents completed a demographic form asking for the age, gender, race, and genetic subtype of their individual with PWS. Parental education and annual family income were also obtained. Demographic variables were used to both describe the sample, and to determine if they related to the FSZ and thus needed to serve as covariates in data analyses.

#### Hyperphagia Questionnaire-Clinical Trial

The HQ-CT was administered to all participants and was used to determine the convergent validity of the FSZ [12]. This 9-item, informant-based questionnaire assesses two components of hyperphagia in PWS; hyperphagic drive or severity, and self-directed food seeking behaviors, and has been used as an endpoint assessing treatment efficacy in clinical trials [13–15].

## Statistical Analyses

### Factor Analyses

Using the combined dataset (n=624), principal component analyses (PCAs) were conducted to identify the latent factor structure of the FSZ. Separate PCA’s were performed using orthogonal (i.e., varimax) versus oblique rotations (i.e., equimax) to assess which rotation yielded the most readily interpretable, conceptually meaningful solutions [25]. Final analyses used the orthogonal solution.

PCAs adhered to well-established criteria [25]. These included Kaiser’s criteria with an eigenvalue > 1; inspection of the Scree Plot to confirm the number of factors; ensuring that items loading onto factors have a common conceptual meaning; nominal cross-loading across factors; factor loadings and communalities >.40; a significant Bartlett’s Test of sphericity; and a Kaiser-Meyer-Olkin measure of sampling adequacy that was close to 1.

Although PCAs were conducted in the follow-up study, the sample size for these analyses was relatively small (n=119), falling below conventional rules of thumb in factor analyses [26]. As such, the goals of these analyses were limited in scope. Specifically, we assessed if the two new items loaded onto factors that made conceptual sense, and if their inclusion compromised the overall structure of the FSZ as established in the large-scale study.

### Internal Consistency

Cronbach’s alphas determined the internal consistency of items within each FSZ factor, and for the overall FSZ in the large-scale study. Alphas were also calculated in the follow-up study to assess the effects of the two additional items on internal consistency.

### Test-Retest Reliability

Test-retest reliability was assessed in 304 participants from the large-scale study who completed the FSZ at time 1 and again 6 months later. To minimize test-retest measurement error, we ensured that raters were the same across assessments. We first compared FSZ scores between Time 1 and Time 2 in matched t-tests. Intraclass Correlation Coefficients (ICCs) were then calculated, which incorporate both the degrees of agreement and correlations between Time 1 and Time 2 [27]. ICCs were based on a single measurement and absolute agreement in a two-way, mixed-effects model in which participant effects were randomized and measure effects were fixed.

### Demographics

Pearson correlations, t-tests or Chi-Squares assessed relations between the FSZ total or factor scores with PWS genetic subtypes, gender, parental education or income, and age. Age was also assessed by dividing participants into three, developmentally appropriate age groups; children (5-12 years), adolescents (13-19 years) and adults (20-59 years; see results).

### Convergent Validity

As food safety tactics are implemented in response to hyperphagia, we predicted that scores on the HQ-CT would be positively associated with the FSZ. Pearson correlations were calculated between the HQ-CT and FSZ total and factor scores. Linear regressions were then conducted with FSZ factors as predictors of the HQ-CT. To further assess relations between the FSZ and hyperphagia, the sample was divided into tertiles based on their total HQ-CT score. ANOVAs then compared FSZ raw factor scores across participants who were in the lowest, middle, and highest tertile HQ-CT groups.

### Open-Ended Question Analyses

A full 78% of parents provided responses to the open-ended question. These comments offer novel insights into how parents implement food-safety practices that complement and extend formal statistical analyses of the FSZ. As such, we conducted a content analysis of their comments, most of which overlapped with items on the FSZ.

## Results

### Factor Analyses

#### Large-Scale Study

Preliminary PCAs with the large, combined dataset revealed that 5 items failed to load onto any factor. These items all dealt with allowing individuals with PWS to attend events with or without food present, and with or without parental supervision. Between 75% to 87% of respondents rated these items “Never,” most likely due to confounds with hyperphagia and intellectual disabilities.

PCAs were then conducted with the remaining 20 FSZ items. One item, “Weigh child at least weekly”, had a poor factor loading (.25) and communality (.35) and was thus eliminated from the final factor analysis. In retrospect, this item did not directly measure food security, and is instead a down-stream indicator of consumed food.

The final PCA with 19 items yielded five factors that collectively accounted for 67.02% of test variance. A common conceptual meaning could be readily applied to these factors, specifically: Alerting Others and Food Supervision in the Community (4 items, 15.32% of rotated variance); Locking and Restricting Food Sources (6 items, 15.21% of variance); Checking for Food (3 items, 14.62% of variance); At Home Supervision and Meals (4 items, 11.80% of variance); and Avoiding Food Settings (2 items, 10.07% of variance).

The Avoiding Food Settings factor contained two items instead of the conventional three or more items. As recommended Worthington and Whittaker [28], we thus ensured that these 2 FSZ items were strongly correlated (r = .72), shared a conceptual meaning, and were relatively unrelated to other FSZ items (r’s ranged from .12 to .31).

Table 2 presents the FSZ items that loaded onto these 5 factors, their factor loadings, and communalities. All factor loadings were strong (or above .40), yet more stringent guidelines suggest that 17 items had either excellent (> .71) or very good (>.63) factor loadings, and just 2 items were deemed fair to good (.40-.45) [29]. Similarly, communalities indicated that all items were valuable in contributing to the test variance of their respective factors. Table 2 also includes the percentages of parents who endorsed each item as all or most of the time versus sometimes or never/rarely.

**Table 2.** FSZ items that loaded onto 5 factors, factor loadings, communalities, item means and relative frequencies from the large-scale-study.

| **FSZ Factor Labels and Items** | **Factor Loading** | **Communalities** | **Item Means (SD)** | **All or Most of the Time** | **Some or None of the Time** |
| --- | --- | --- | --- | --- | --- |
| **1. Alert Others, Supervision in the Community** |  |  |  |  |  |
| Make sure adults involved with my child are aware of his/her food issues. | .79 | .66 | 3.68 (.74) | 91.7% | 8.3% |
| Alert others of child's food issues (to ensure they do not give child access to food). | .78 | .67 | 3.47 (.91) | 85.0% | 15.0% |
| Make sure they are supervised while away from home. | .76 | .66 | 3.64 (.75) | 89.7% | 10.3% |
| Ensure child has no access to other people's food at school, camp, or work. | .67 | .53 | 3.06 (1.16) | 72.1% | 27.9% |
| Make a food plan for child prior to attending events, outings, restaurants* | .62 | .56 | 3.24 (.89) | 79.7% | 20.3% |
| **2. Lock, Restrict Food Sources** |  |  |  |  |  |
| Lock up pantry or cabinets where food is kept | .90 | .87 | 2.64 (1.35) | 55.3% | 44.7% |
| Lock up refrigerator or freezer | .89 | .80 | 2.46 (1.40) | 49.9% | 50.1% |
| Ensure there is no food left on counter tops, tables, or other areas of access | .65 | .68 | 2.99 (1.35) | 70.2% | 29.8% |
| Lock up trash, compost, or recycling bins | .50 | .51 | 1.62 (1.35) | 20.5% | 79.5% |
| Keep money or credit cards from child | .46 | .50 | 2.32 (1.36) | 46.0% | 54.0% |
| Use security features (alarm, camera) in home to monitor food access | .40 | .47 | 1.62 (1.23) | 20.7% | 79.3% |
| **3. Check for Food** |  |  |  |  |  |
| Check their person (pockets, pat down, shoes, etc.) for food, wrappers, or money | .87 | .82 | 1.70<br>(.94) | 18.6% | 81.4% |
| Check their belongings or bedroom for food, wrappers, or money | .87 | .83 | 1.98<br>(1.03) | 28.0% | 72.0% |
| Check on child in bathroom to be sure he or she is not eating food | .84 | .75 | 1.69<br>(.99) | 19.6% | 80.4% |
| **4. At Home Supervision, Meals** |  |  |  |  |  |
| Make sure child is busy while at home | .72 | .66 | 2.86<br>(.85) | 67.6% | 32.4% |
| Avoid eating in front of child unless they are also eating* | .69 | .55 | 2.86<br>(1.02) | 67.0% | 33.0% |
| Aware of where child is at home all the time (but may not be in sight of caregiver) | .63 | .63 | 3.39<br>(.86) | 85.6% | 14.4% |
| Supervise - always have eyes on at home | .61 | .62 | 3.02<br>(.93) | 73.9% | 26.1% |
| Make sure that meals are planned and on time | .50 | .41 | 3.05<br>(.81) | 79.4% | 20.6% |
| **5. Avoid Food Settings** |  |  |  |  |  |
| Avoid taking child to the grocery store | .85 | .79 | 2.08<br>(1.07) | 34.3% | 65.7% |
| Avoid taking child to restaurants | .84 | .80 | 2.18<br>(1.02) | 34.3% | 60.2% |

**Notes:** \*Shaded items are from the follow-up study and included here for reader convenience. See S1 Table for the complete PCA results from the follow-up study.

#### Follow-Up Study

PCA’s conducted in the follow-up study assessed the impact of the 2 additional items on the overall structure of the FSZ. The PCA with varimax rotation yielded 5 factors that collectively accounted for 62.75% of test variance. These 5 factors recapitulated the factors derived in the large-scale study, albeit with slightly different factor loadings, communalities, and frequencies (see S1 Table 1). Importantly, the two new items loaded onto factors that were readily interpretable. The item “Make a food plan for child prior to attending events, outings, restaurants” loaded onto the Alerting Others and Supervision in the Community factor, and “Avoid eating in front of child unless they are also eating” was consistent with other tactics families implement in the At Home Supervision and Meals factor. The two added items were frequently endorsed, 79.7% and 67%, respectively. T-tests comparing all FSZ factor mean scores between the large-scale and follow-up studies were not significant (see S2 Table).

### Internal Consistency

Cronbach’s alphas were calculated for each of the five factors derived from the large-scale and follow-up studies. Conventional rules of thumb suggest that alphas >.70 and < .90 are considered good [30]. As summarized in Table 3, all alphas fell into this range and varied from .73 to .89.

**Table 3.** Cronbach alphas for FSZ factors in both the large-scale and follow-up studies.

| FSZ Factors | Cronbach Alpha's |
| --- | --- |
| Alert Others, Supervise in Community Large-Scale Study | .79 |
| Alert Others, Supervise in Community Follow-up Study | .76 |
| Lock, Restrict Food Sources Large-Scale Study | .82 |
| Lock, Restrict Food Sources Follow-up Study | .79 |
| Check for Food Large Scale Study | .89 |
| Check for Food Follow-Up Study | .89 |
| At Home Supervision, Meals Large-Scale Study | .76 |
| At Home Supervision, Meals Follow-Up Study | .73 |
| Avoid Food Settings Large-Scale Study | .81 |
| Avoid Food Settings Follow-Up Study | .74 |
| Total FSZ Raw Scores Large Scale Study | .89 |
| Total FSZ Raw Scores Follow-Up Study | .86 |

### Test-Retest Reliability

Matched t-tests conducted with the total FSZ scores between Time 1 and Time 2 proved nonsignificant, Time 1 <u>M</u> = 51.93, <u>SD</u> = 13.07, Time 2 <u>M</u> = 51.62, <u>SD</u> = 12.65. Intraclass correlations and their corresponding 95% confidence intervals are presented in Table 4. Based on conventional criteria, ICCs were all in the good range [31].

**Table 4.** FSZ Time 1 to Time 2 Intraclass Correlations and 95% Confidence Intervals.

| FSZ Factors | Intraclass Correlations (95% CI) |
| --- | --- |
| Alert Others, Supervision in the Community | .74 (.67-.78) |
| Lock, Restrict Food Sources | .88 (.86-.91) |
| Check for Food | .78 (.74-.82) |
| At Home Supervision, Meals | .75 (.69-.79) |
| Avoid Food Settings | .65 (.58-.71) |
| Total FSZ | .88 (.85-.90) |

### Combined Large-Scale and Follow-Up Studies

Separate analyses of the large-scale and follow-up studies yielded the same factor structure, as well as similar mean FSZ scores and Cronbach’s alphas. As such, these datasets were combined (n = 743) for subsequent analyses of relations between the FSZ and demographic variables, as well as the convergent validity of the FSZ.

### FSZ and Demographics

ANOVAS or t-tests revealed no significant differences in FSZ total or factor scores across gender, PWS genetic subtypes, race, and parental income or education. The correlation between age and total FSZ scores was relatively small, <u>r</u> (733) = .19, <u>p</u>< .001. Correlations also assessed relations between age and the HQ-CT’s total score, and the severity/drive and food-seeking behavior domains. Only the self-directed food-seeking behavior domain proved significant, <u>r</u> (733) = .22, <u>p</u><.001.

To further explore these findings, participants were divided into three developmentally appropriate age groups: children aged 5 through 12 years (n = 268; <u>M</u> = 8.31 years, <u>SD</u> = 2.44); adolescents aged 13-19 years (n=247, <u>M</u> = 16.16 years, <u>SD</u> = 2.40) and adults aged 20-59 years (n=228, <u>M</u> = 30.50 years, <u>SD</u> = 8.36). Between-age group ANOVAs were then conducted with both FSZ and HQ-CT.

As summarized in Table 5, significant age group differences were found in 4 FSZ factors. Bonferroni post-hocs revealed that locking food sources differed significantly between all age groups, and that the adolescent and adult groups scored higher than children in checking for food and avoiding food setting. Children, however, scored higher than the 2 older age groups in the alerting others factor. Effect sizes (η^2^) were large for the locking factor, medium for checking for food, and small for the remaining two factors.

**Table 5.**
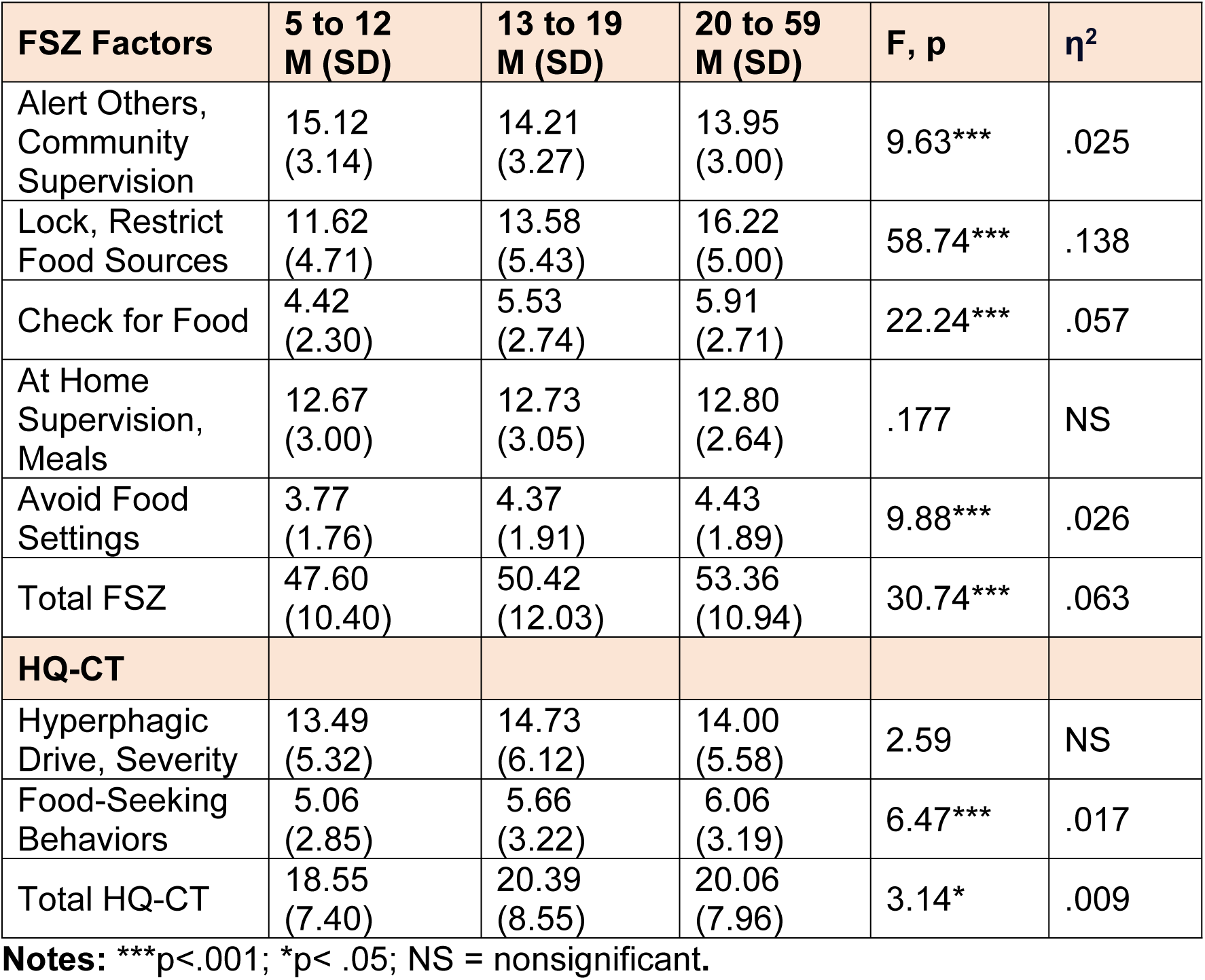
FSZ and HQ-CT mean raw scores, and F, p and η^2^ values across three age groups.

| FSZ Factors | 5 to 12<br>M (SD) | 13 to 19<br>M (SD) | 20 to 59<br>M (SD) | F, p | $\eta^2$ |
| --- | --- | --- | --- | --- | --- |
| Alert Others,<br>Community<br>Supervision | 15.12<br>(3.14) | 14.21<br>(3.27) | 13.95<br>(3.00) | 9.63*** | .025 |
| Lock, Restrict<br>Food Sources | 11.62<br>(4.71) | 13.58<br>(5.43) | 16.22<br>(5.00) | 58.74*** | .138 |
| Check for Food | 4.42<br>(2.30) | 5.53<br>(2.74) | 5.91<br>(2.71) | 22.24*** | .057 |
| At Home<br>Supervision,<br>Meals | 12.67<br>(3.00) | 12.73<br>(3.05) | 12.80<br>(2.64) | .177 | NS |
| Avoid Food<br>Settings | 3.77<br>(1.76) | 4.37<br>(1.91) | 4.43<br>(1.89) | 9.88*** | .026 |
| Total FSZ | 47.60<br>(10.40) | 50.42<br>(12.03) | 53.36<br>(10.94) | 30.74*** | .063 |
| **HQ-CT** |  |  |  |  |  |
| Hyperphagic<br>Drive, Severity | 13.49<br>(5.32) | 14.73<br>(6.12) | 14.00<br>(5.58) | 2.59 | NS |
| Food-Seeking<br>Behaviors | 5.06<br>(2.85) | 5.66<br>(3.22) | 6.06<br>(3.19) | 6.47*** | .017 |
| Total HQ-CT | 18.55<br>(7.40) | 20.39<br>(8.55) | 20.06<br>(7.96) | 3.14* | .009 |

**Notes:** \*\*\* $p < .001$ ; \* $p < .05$ ; NS = nonsignificant.

As shown in Table 5 the ANOVA assessing age group differences in the total HQ-CT was marginally significant. Consistent with correlational analyses, this finding is driven by the more robust age group differences in the HQ-CT’s food seeking domain.

### Convergent Validity

As expected, total FSZ and HQ-CT scores were positively correlated *r* (725) = .54, <u>p</u>< .001. Indeed, total HQ-CT scores were significantly correlated with all 5 FSZ factors, (r’s range =.29 to .50, <u>p</u>’s< .001).

The regression predicting the total HQ-CT was significant, <u>F</u> (5,717) = 66.13, <u>p</u> < .001, adjusted R^2^ = .324. FSZ predictors, with <u>p</u>’s < .001, included Checking for Food, *t* =11.19, β = .401; Locking Food Sources, *t* = 6.30, β = .256; and At Home Supervision and Meals, *t* = 3.21, β = .153. Avoiding Food Settings was marginally significant, *t* = 2.33, <u>p</u> = .020, β = .082.

Probing deeper, participants were categorized into tertiles based on their total HQ-CT raw score; ANOVAs then compared FSZ scores across hyperphagia tertiles. The lowest tertile included 238 participants (32%), the middle tertile contained 245 individuals (33%) and the highest group had 260 participants (35%). As the tertile groups did not significantly differ in age, age was not controlled for in ANOVAs. As summarized in Table 6, significant differences were found between tertiles and all five of the FSZ raw factor scores. Bonferroni post-hocs revealed that all groups differed significantly from one another, with one exception. The medium and highest tertiles had comparable scores on the At Home Supervision and Meals factor. Effect sizes (η^2^) were large for 3 factors, and medium to large for 2 factors.

**Table 6.**
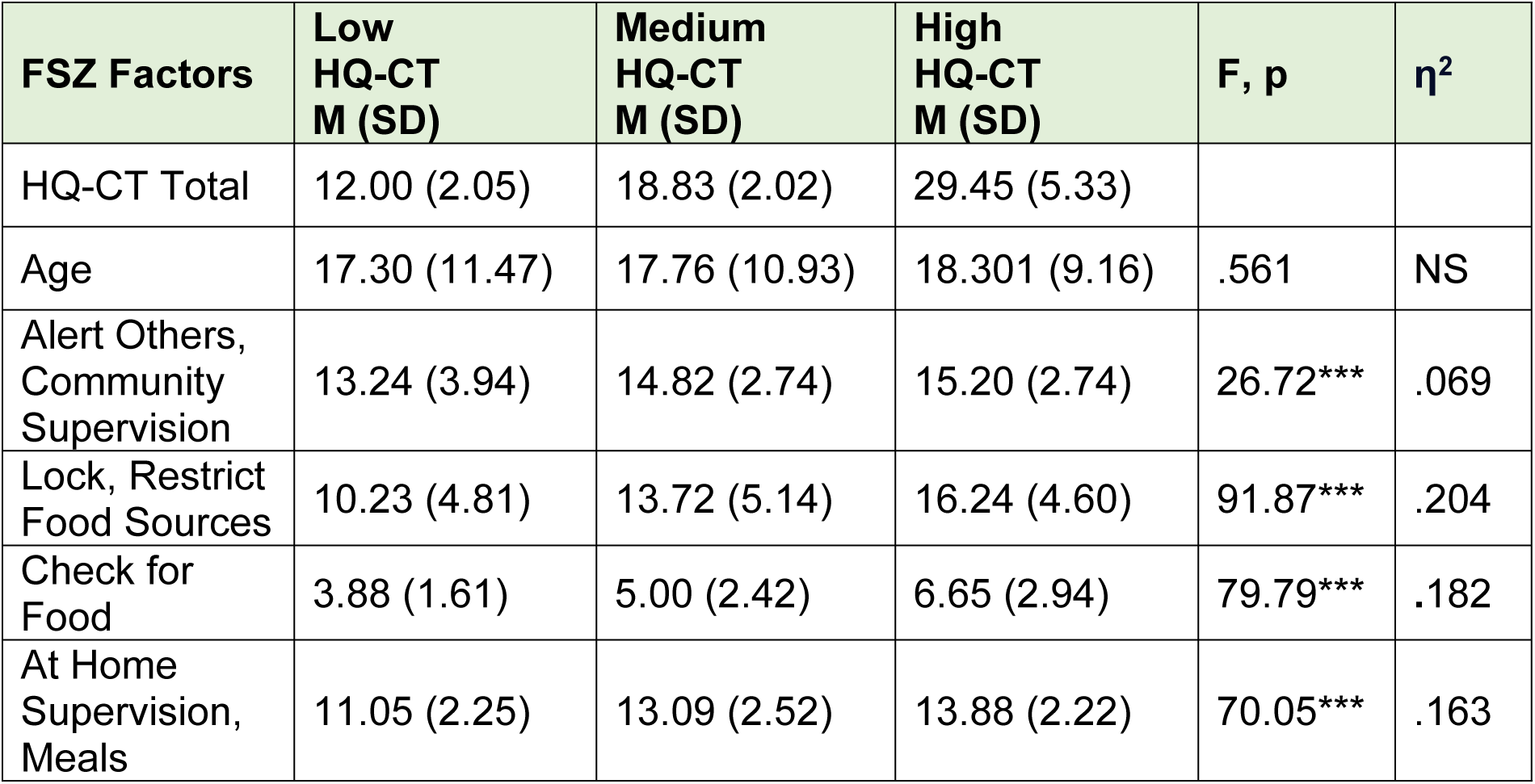
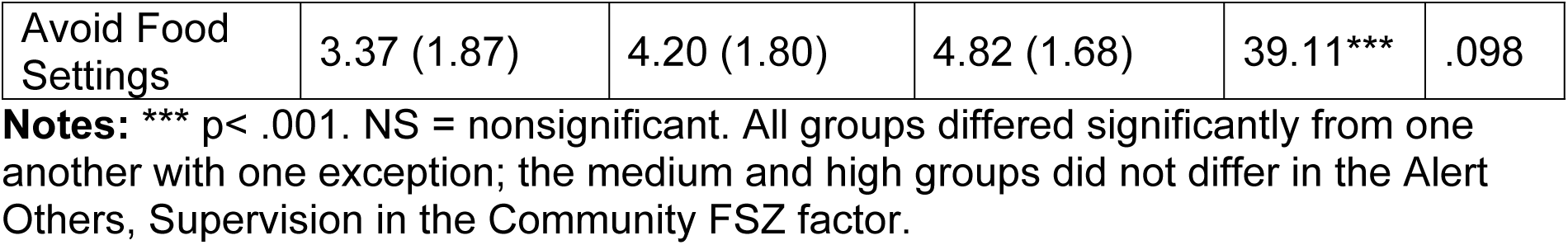
Comparisons of FSZ raw factor scores across tertiles of participants with low, medium, and high total HQ-CT scores.

| FSZ Factors | Low<br>HQ-CT<br>M (SD) | Medium<br>HQ-CT<br>M (SD) | High<br>HQ-CT<br>M (SD) | F, p | $\eta^2$ |
| --- | --- | --- | --- | --- | --- |
| HQ-CT Total | 12.00 (2.05) | 18.83 (2.02) | 29.45 (5.33) |  |  |
| Age | 17.30 (11.47) | 17.76 (10.93) | 18.301 (9.16) | .561 | NS |
| Alert Others,<br>Community<br>Supervision | 13.24 (3.94) | 14.82 (2.74) | 15.20 (2.74) | 26.72*** | .069 |
| Lock, Restrict<br>Food Sources | 10.23 (4.81) | 13.72 (5.14) | 16.24 (4.60) | 91.87*** | .204 |
| Check for<br>Food | 3.88 (1.61) | 5.00 (2.42) | 6.65 (2.94) | 79.79*** | .182 |
| At Home<br>Supervision,<br>Meals | 11.05 (2.25) | 13.09 (2.52) | 13.88 (2.22) | 70.05*** | .163 |
| Avoid Food Settings | 3.37 (1.87) | 4.20 (1.80) | 4.82 (1.68) | 39.11*** | .098 |

**Notes:** \*\*\* $p < .001$ . NS = nonsignificant. All groups differed significantly from one another with one exception; the medium and high groups did not differ in the Alert Others, Supervision in the Community FSZ factor.

### Analyses of Open-Ended Question Responses

Beyond the two new items, four sets of findings also emerged from the open-ended question. First, 21 respondents indicated that their individual with PWS did not engage in food-seeking behaviors. The majority of these (n=16, 76.2%) were noted by their parents to have a medical or psychiatric exceptionality that impacted their hyperphagia or food intake. These exceptionalities were: severe developmental delays, including four, minimally verbal individuals with autism spectrum disorder, and two individuals with ongoing psychotic episodes. The remainder had such medical complications as Type I diabetes (n=3), multiple food allergies with anaphylaxis (n=3), severe hypothermia (n=1), overwhelming fatigue (n=1), being G-Tube dependent (n=1) and being paralyzed and in a wheelchair (n=1). We were curious if eliminating these individuals would substantially shift PCA or other analyses; it did not.

Second, parents overwhelmingly responded to the open-ended question by elaborating on the specific ways that they individualized or implemented FSZ tactics. Our team reviewed these descriptions and readily classified them into 6 categories that dovetailed with FSZ items, specifically: locking and securing food sources; scheduling meals and snacks; managing restaurants, parties, and family gatherings; eating and discarding food at home; and working with schools. Table 7 summarizes examples of parental responses within these categories, as well as the frequencies of them relative to the number of respondents. As parents could elaborate on several strategies the frequencies noted in Table 9 exceeds 100%.

**Table 7.**
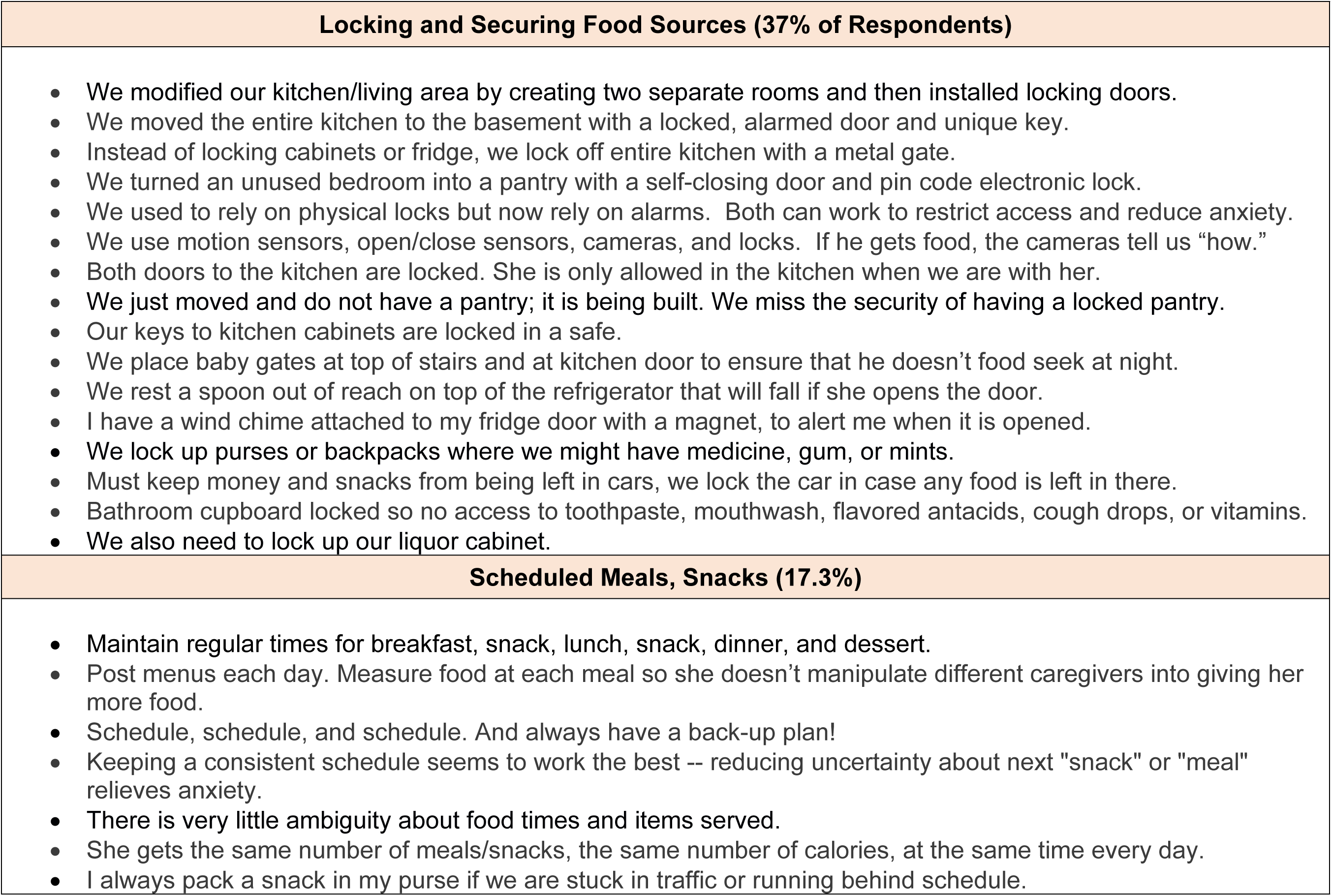
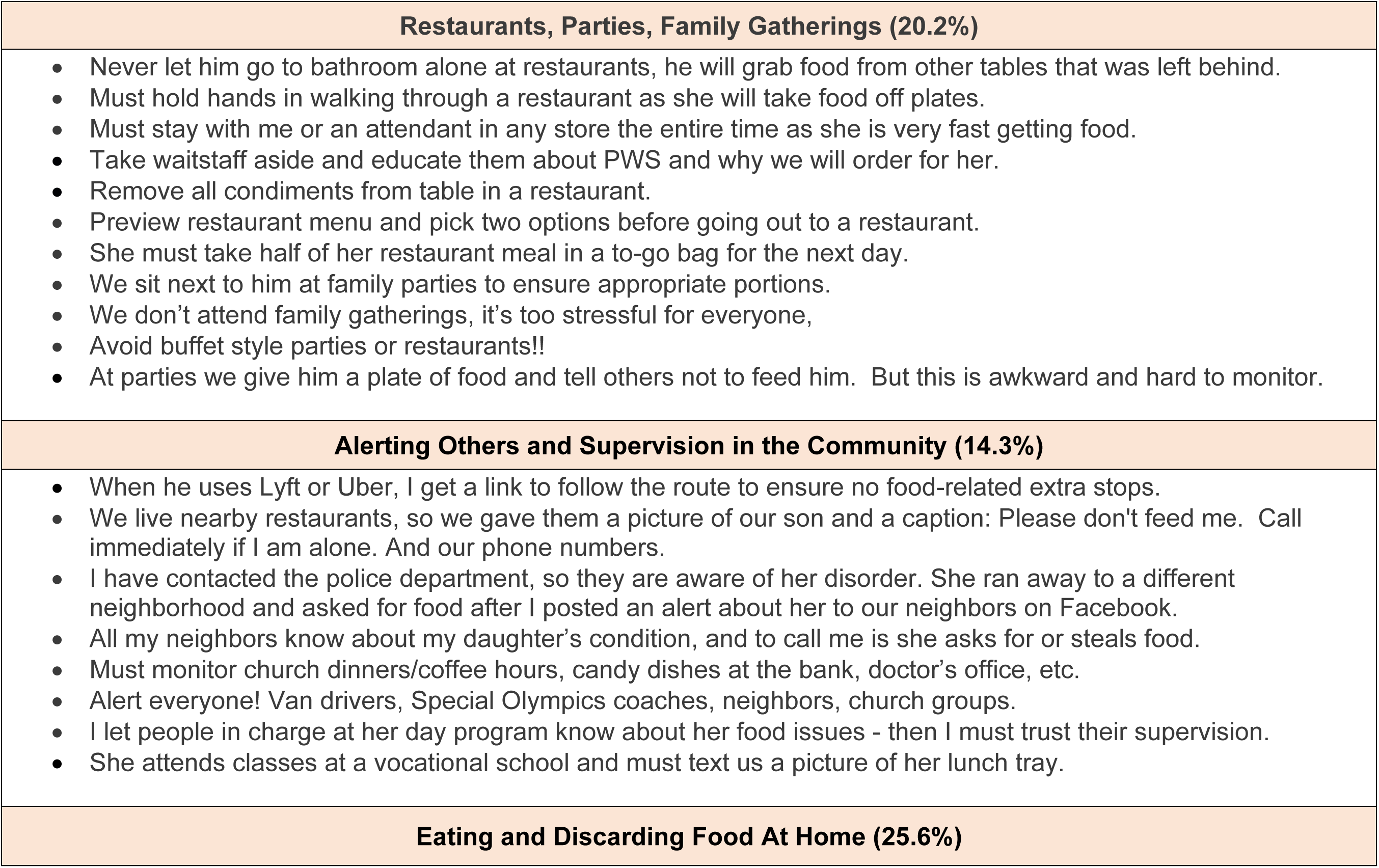
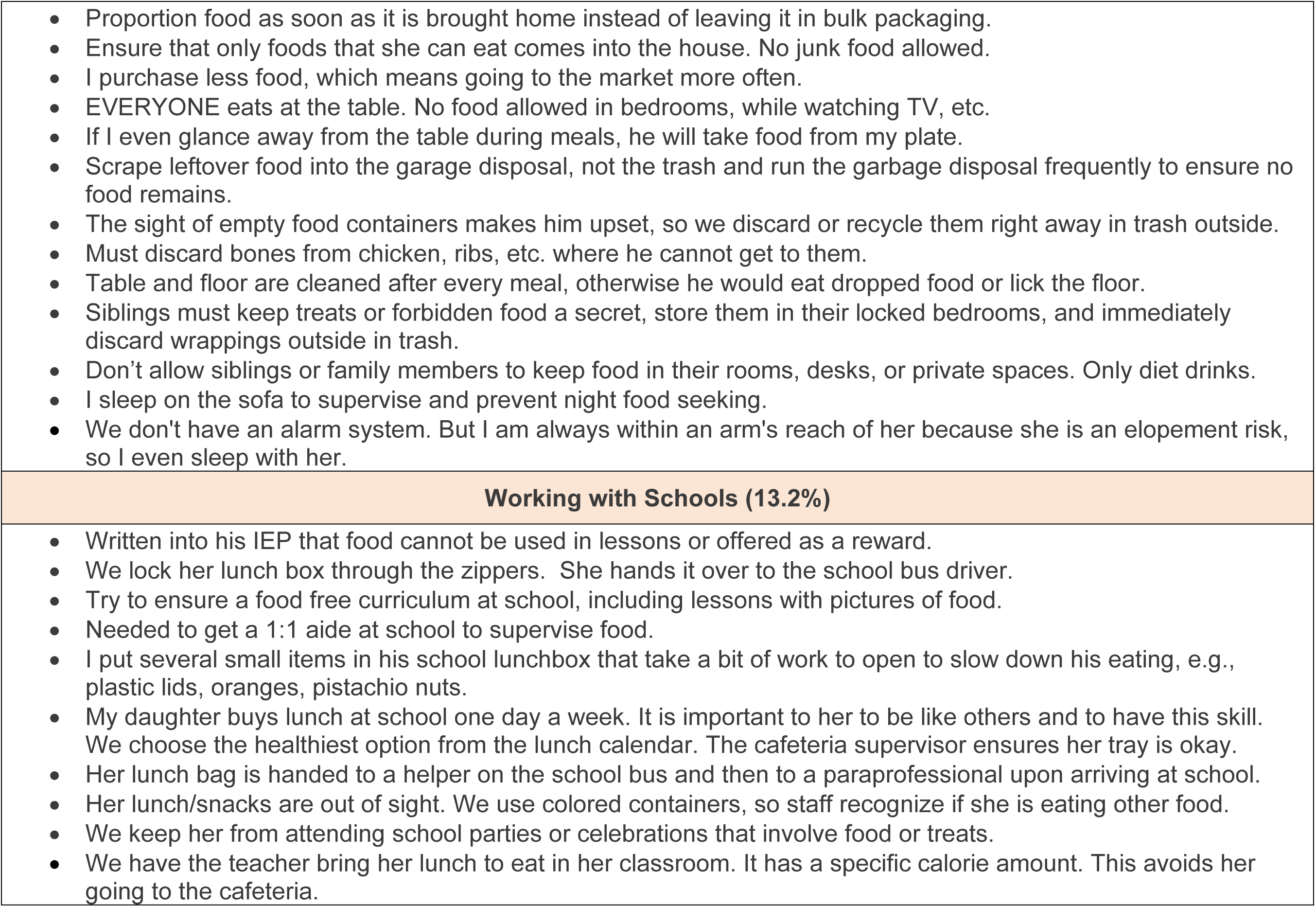
Examples and frequencies of parental descriptions of food FSZ tactics derived from the open-ended question.

Third, 9.1% of respondents offered ways that they help their individual with PWS to eat. Although not necessarily food security tactics, parents noted that they used smaller plates so that food quantities seem larger, pre-plated meals, cut-up portions to avoid overstuffing, and encouraged their child to put down their fork after every few mouthfuls. Several participants also emphasized the importance of teaching their individual with PWS about proper nutrition and establishing healthy eating habits in all family members.

Finally, 5 respondents offered that their individual with PWS consumed non-edible items (e.g., pieces of a TV remote, hair). Some people with PWS may eat unpalatable items (e.g., stick of butter, frozen meat, garbage, pet food), or endorse a willingness to eat unusual food combinations [32]. However, pica, or eating non-food items, has not been widely reported in the PWS literature.

## Discussion

The FSZ emerged as a psychometrically sound index of the tactics that parents use to manage their child’s hyperphagia that stands to enhance their accurate assessments of hyperphagia in future research or clinical trials. Beyond the psychometrics of the FSZ, findings are discussed in relation to hyperphagia, and how food safety tactics compare to family accommodation in other disorders. As well, we discuss how parents tailor their food safety tactics to meet the individual needs of their child with PWS, and the impact of doing so on their well-being.

Consistent with best practices in creating novel questionnaires [22], the FSZ was developed with input from multiple stakeholders, including parents, PWS specialists and researchers, and individuals with PWS. In an iterative feedback process, items were added, or revised for clarity, and then pilot-tested, revised, and administered to parents in a large-scale study. Based on parental responses to an open-ended question, two additional items were added, and subsequently evaluated in a follow-up study. This multi-step process helped ensure both the construct and content validity of the FSZ.

Other psychometric properties of the FSZ were also robust. PCAs in the large-scale study yielded five, conceptually meaningful factors that collectively accounted for 67% of test variance. Communalities indicated that all items contributed meaningfully to their respective factors. Cronbach’s alphas revealed strong internal consistency of items within each factor, and for FSZ total scores. As well, ICC’s suggest strong test-retest reliability. Indeed, mean FSZ scores at Time 1 and 2 were almost identical, suggesting relatively stability in FSZ tactics over this 6-month time interval.

Importantly, analyses in the follow-up study, with two additional items, revealed the same overall factor structure as the large-scale study, albeit with slight differences in factor loadings, communalities, and frequencies. The new items were frequently endorsed by respondents and loaded onto two factors that made conceptual sense. Further, no differences were found between the large-scale and follow-up studies in mean scores of any FSZ factors. Taken together, findings justify the use of the final, 21-item version of the FSZ questionnaire in future research or clinical trials.

Age was the only demographic variable significantly associated with the FSZ. Relative to children, parents of adolescents and adults were more apt to lock food sources, check for food and avoid food settings. Parents of children, however, scored higher than their counterparts in the alerting others about food issues. These findings are best understood in relation to significant age-related increases in the HQ-CT’s food-seeking behavior domain, even as hyperphagic severity or drive remained relatively stable in participants.

With advancing age and development, people with PWS may become more skilled or adept in finding or sneaking food. Indeed, they are known to exhibit such ingenious food-seeking strategies as unscrewing hinges to kitchen cabinets at night, dangling food on strings in the heating vents, and memorizing credit card numbers and ordering food deliveries to a friend’s address. Such tactics require foresight and planning yet contradict well-documented deficits in executive functioning in people with PWS, especially task-switching and planning abilities [ 7, 8]. Perhaps these contradictory findings can be partially explained by hunger. Al-Shawaf [33] reports that people in states of chronic or acute hunger have difficulties sustaining attention on food-irrelevant tasks, thereby compromising their general planning and problem-solving abilities. At the same time, however, hunger enhances memory of food stimuli [34] and the ability to solve food-acquisition problems [33].

Second, transitioning from childhood into adolescence or adulthood typically brings more opportunities for individuals to engage in community activities outside of the family home. And, compared to home, food is apt to be more readily available in community recreational, educational, or vocational venues. As one parent noted “My 23-year-old is more independent now, and he has found a church down the street that feeds him.”

Convergent validity analyses of the FSZ confirmed the hypothesized relationship between the FSZ and hyperphagia. Regression analyses revealed that all but one FSZ factor was predictive of the total HQ-CT. Follow-up comparisons of FSZ factors across those with low, medium, or high HQ-CT scores revealed robust differences in the expected direction in all five FSZ factors. These data offer a point of reference for using the FSZ in future studies or clinical trials. Importantly, age did not differ across HQ-CT tertiles, indicating that parents implement FSZ strategies in response to their child’s hyperphagic symptoms, not necessarily their age.

One FSZ factor was not a significant predictor of hyperphagia, Alerting Others and Community Supervision. Even so, all items in this factor were frequently endorsed, especially in the youngest age group. It is possible that, given their child’s PWS diagnosis, families preemptively alert others about their child’s food issues as a baseline strategy. This widespread strategy remains in place, even as parents implement additional FSZ tactics in response to the changing needs of their individual with PWS.

The FSZ is a means of reminding parents of the many ways that they accommodate to their child’s hyperphagia. A related literature examines accommodation in parents of children with psychiatric conditions, especially anxiety disorders [35, 36]. In these families, accommodation refers to how parents modify their behaviors to alleviate or avoid the distress, anxiety or maladaptive behaviors caused by their child’s disorder. In PWS, food safety tactics also alleviate anxiety, including food-related temper outbursts or repetitive questioning [4].

Although parental accommodation in both instances may temporarily reduce child anxiety, they lead to markedly different outcomes. Higher levels of parental accommodation to their children’s psychiatric symptoms backfires, and in the long-term is associated with increased symptomatology, impairment, and poorer responses to treatment [35, 36]. In contrast, food safety tactics in PWS are lifesaving. In the long-term, however, food safety practices may become the “new normal,” potentially compromising accurate parental evaluations of hyperphagia.

Parental responses to the open-ended question offered poignant insights into both the logistics and stress of ensuring food safety. Several overarching messages emerged from their comments. First, given the high response rate to this question, parents were clearly motivated to explain their food safety practices. Similarly, parents in the focus groups we conducted in developing the FSZ offered that they are rarely asked to discuss food safety and welcomed the opportunity to do so.

Second, parental remarks highlighted that food safety is a life-long, round the clock pursuit, one that is especially challenging given the necessity for humans to eat and the omnipresence of food in social and community settings. And food safety requires constant vigilance. As one parent offered, “If the food is not secured and she gets it, then it is our fault. Not hers. This syndrome is terrible for them to live with.”

Third, parents were incredibly diligent and creative in establishing food security tactics at home, school, and in the community. At the same time, however, these parents also experience markedly high levels of caregiving burden. Indeed, levels of caregiving burden in PWS are high even as compared to parents of children with autism spectrum disorder or older caregivers of spouses with dementia [37]. Such burden relates to managing both their child’s hyperphagia and behavior problems, and has a profound, negative impact on their social and personal quality of life [37]. As one mother offered “My ENTIRE life is food security. 24/7, 24/7. We have NO life!”

Added to this burden is a counterintuitive psychological dilemma—the parental instinct to nurture and feed their hungry children juxtaposed with the reality that doing so could compromise their health and longevity. As one participant astutely observed, “It is so hard to balance the psychology of the parent, of wanting to feed a hungry child, with the dire medical repercussions of doing so.”

Finally, and as Table 7 depicted, there is no single “right” way of practicing food security. Some parents, for example, avoid attending restaurants or social gatherings, others find ways of navigating them. Some keep their children away from the school cafeteria, others do not. Some allow siblings to keep treats locked in their bedrooms, others forbid it. Some parents use locks and alarms, others do not. Parents are thus implementing FSZ tactics that meet the individual and changing needs of their child with PWS while also considering what is feasible within the larger context of their family.

Several study weaknesses should be noted. First, participants were generally White, well educated, and with relatively high annual incomes. Although SES was not significantly related to the FSZ, it would be helpful for future studies to include more economically or racially diverse participants. In this vein, one respondent offered that being homeless and in temporary housing made food safety nearly impossible. Second, test-retest reliability is typically assessed across shorter time frames than the 6-month interval used in the current study. Even so, mean FSZ scores at Time 1 and 2 were almost identical, and Intra-class correlations were strong. Finally, we did not examine associations between the FSZ and family composition or marital status. Future research is needed on how, for example, siblings, grandparents, current or ex-spouses practice or perceive food safety.

Despite these concerns, this study is the first to document and analyze food safety tactics in PWS. The 21-item FSZ emerged as a psychometrically robust measure of parental food safety tactics aimed at enhancing their accurate evaluations of hyperphagia in future research or clinical trials. In the meantime, the study also shines a light on the never-ending and extraordinary measures that parents use to ensure the health and wellbeing of their loved one with PWS.

## Data Availability

All relevant data are within the manuscript and its Supporting Information files.

## Acknowledgements

The authors thank the many parents who participated in this study, as well as the generosity of stakeholders in providing their time and essential feedback in developing the FSZ. We are especially grateful for the enthusiastic support of our research from the Foundation for Prader-Willi Research (FPWR), including Teresa Strong PhD, Lauren Schwartz PhD, Jessica Bohonowych PhD, and members of the FPWR Clinical Trials Consortium. The authors also thank Robert M Hodapp, PhD, for his critical reading of an earlier draft of this manuscript.

## Notes

### Competing Interest Statement

The authors have declared no competing interest.

### Funding Statement

The author(s) received no specific funding for this work.

